# Post-traumatic stress symptoms in cancer patients during the COVID-19 pandemic: a one-year longitudinal study

**DOI:** 10.1101/2022.01.11.22269053

**Authors:** Etienne Bastien, Sophie Lefèvre-Arbogast, Justine Lequesne, François Gernier, François Cherifi, Olivier Rigal, Lydia Guittet, Jean-Michel Grellard, Giulia Binarelli, Marie Lange, Marie Fernette, Laure Tron, Adeline Morel, Doriane Richard, Bénédicte Griffon, Alexandra Leconte, Florian Quilan, Louis-Ferdinand Pépin, Fabrice Jardin, Marianne Leheurteur, Audrey Faveyrial, Bénédicte Clarisse, Florence Joly

**Affiliations:** Medical Oncology Department, François Baclesse Center, Caen France; Anticipe (Interdisciplinary Research Unit for the Prevention and Treatment of Cancer), INSERM Unit 1086, Caen France; Clinical Research, François Baclesse Center, Caen France; Medical Oncology, Henri Becquerel Center, Rouen France; Clinical Research, Henri Becquerel Center, Rouen France; Hematology, Henri Becquerel Center, Rouen France

## Abstract

**Background:** Cancer patients may be particularly vulnerable to psychological consequences of the COVID-19 pandemic and successive lockdowns. We studied the prevalence and evolution of post-traumatic stress disorder (PTSD) symptoms in cancer patients during the pandemic waves, and investigated factors associated with high symptoms.

**Methods:** COVIPACT is a one-year longitudinal prospective study of French patients with solid/hematologic malignancy receiving treatment during the first nationwide lockdown. PTSD symptoms were measured every 3 months from April 2020 using the Impact of Event Scale-Revised. Patients also completed validated questionnaires on quality of life (QoL), cognitive complaints and insomnia, and a survey on their COVID-19 lockdown experience.

**Results:** Longitudinal analyses involved 386 patients with at least one PTSD assessment after baseline (median age 63, 76% female). Among them, 21.5% had moderate/severe PTSD symptoms during the first lockdown. The rate of patients reporting PTSD symptoms decreased at lockdown release (13.6%), increased again at second lockdown (23.2%), and slightly declined from the second release period (22.7%) to the third lockdown (17.5%). Patients were grouped into three trajectories of evolution. Most patients had stable low symptoms throughout the period, 6% had high baseline symptoms slowly decreasing over time, and 17.6% had moderate symptoms worsening during second lockdown. Female sex, feeling socially isolated, worrying about COVID-19 infection, and using psychotropic drugs were associated with PTSD symptoms. PTSD symptoms were associated with impaired QoL, sleep and cognition.

**Conclusions:** Around a quarter of cancer patients presented high and persistent PTSD symptoms over the first year of the COVID-19 pandemic and may benefit from psychological support.

## INTRODUCTION

The COVID-19 pandemic spread worldwide from China in January 2020 and led to national measures such as lockdowns and curfews in many parts of the world as soon as March/April 2020. This stressful event has deeply and lastingly impacted populations in health, psychological, social and financial terms.^1,2^ Post-traumatic stress disorder (PTSD) is defined as the direct or indirect exposure to a stressor (*e.g*., actual or threatened death) followed by symptoms of intrusive memories, avoidance and hyperarousal that persist over one month.^3^ PTSD and related disorders like anxiety, distress and depression have been reported during past pandemics such as Ebola, severe acute respiratory syndrome (SARS) and H1N1 influenza, and their effects are compounded by the public health measures required, such as quarantine.^4^ The COVID-19 pandemic is unprecedented owing to the repetition and duration of multiple waves of infections and lockdowns and may have deeper, long-term psychological consequences.

Some studies have reported high levels of PTSD symptoms in the general population during the COVID-19 pandemic, although most of them were cross-sectional or with short-term follow-up. A meta-analysis including 39 studies in the general population estimated a prevalence of PTSD symptoms of 27% during the first wave of the pandemic.^5^ Very few studies repeated these assessments to investigate changes in PTSD over time. Wang *et al*. assessed the PTSD symptomatology of a sample of Chinese individuals twice during the first wave of the pandemic.^6^ They observed high levels of PTSD symptoms during China’s COVID-19 outbreak (end of January 2020), without any clinically significant change one month later. In a longitudinal French study, 35.5% of the population showed clinically significant COVID-19 peritraumatic distress during the first lockdown (April 2020) compared to 17.2% three months later after the lifting of restrictions (July 2020).^7^ This distress was associated with being female, a student, having pre-existing mental health problems and higher levels of worries about the COVID-19 crisis.

Cancer patients may be particularly prone to develop PTSD symptoms due to a high baseline level of anxiety and depression in relation with their cancer diagnosis,^8^ the potential modification of cancer management in response to the COVID-19 pandemic^9^, and a high risk of developing a severe form of COVID-19 infection.^10–12^ Several studies in various types of cancer reported PTSD symptoms in patients during the first wave of the pandemic, with a prevalence ranging from 9% to 35%.^13–15^ However, to our knowledge, there has been no longitudinal study to date following cancer patients over the different waves of the pandemic.

As soon as April 2020, we set up a one-year longitudinal prospective clinical cohort of cancer patients (COVIPACT). Baseline results showed that 21% of cancer patients experienced PTSD symptoms associated with poor quality of life (QoL) during the first lockdown.^16^ An adjustment to medical oncology practices was implemented in approximately one-quarter of patients and was associated with the occurrence of PTSD symptoms. We present here the longitudinal follow-up of the COVIPACT cohort. Our main objective was to describe the evolution of PTSD symptoms in cancer patients over the first year of the COVID-19 pandemic and to identify factors associated with high PTSD symptoms. We also investigated the evolution of insomnia, cognitive complaints and QoL over the pandemic in relation to PTSD symptoms.

## METHODS

### Settings and population

COVIPACT (NCT104366154) is a French longitudinal prospective study investigating the psychological impact of the COVID-19 pandemic on cancer patients. The study included outpatients aged 18 and over, and receiving oncological treatment for solid or hematological malignancy during the first nationwide lockdown at the day care hospital of two French cancer centers (the François Baclesse Center and the Henri Becquerel Center).^16^ Patients were enrolled between April, 16, 2020 and May, 29, 2020.

### Data collection and survey

Demographics, clinical information, history of oncological disease and pandemic-induced adjustment in medical oncology practices were collected on medical records. At baseline, enrolled patients were asked to complete a set of validated self-questionnaires assessing PTSD symptoms, insomnia, QoL and cognitive complaints. Follow-up questionnaires were then sent by mail every 3 months over a year, which approximately matched the waves of the pandemic and related restrictions. Questionnaires were completed in April-May 2020 during the first French lockdown (M0), in July-August 2020 during the release period (M3), in October-November 2020 during the second French lockdown (M6), in January-February 2021 after the second lockdown release (M9), and in April-May 2021 during the third French lockdown. Patients were included in the present study if they completed the questionnaire on PTSD symptoms at baseline and at least one follow-up.

#### Assessment of post-traumatic stress symptoms

The Impact of Event Scale-Revised (IES-R) is a 22-item self-questionnaire which highlights avoidance, intrusion, and hyperarousal PTSD symptoms in relation to a pre-specified stressful event.^17,18^ As part of the COVIPACT study, patients were asked to complete the IES-R questionnaire as regards the COVID-19 pandemic. As previously recommended, a score of 33 or higher out of 88 was used to identify moderate-to-severe PTSD symptoms.^18^

#### Assessment of quality of life

We used the Functional Assessment of Cancer Therapy - General (FACT-G) to assess QoL.^19^ It includes 27 items that measure the physical, social, emotional and functional components of health-related QoL in cancer patients and yields a total score of 108.

#### Assessment of insomnia

Insomnia was assessed by the Insomnia Severity Index (ISI).^20^ This short 7-item scale takes into account night-time and daytime symptoms of insomnia. The total score ranges from 0 to 28 and is interpreted as follows: absence of insomnia (0–7); sub-threshold insomnia (8– 14); moderate insomnia (15–21); and severe insomnia (22–28).

#### Assessment of cognitive complaints

Cognitive complaints were evaluated by The Functional Assessment of Cancer Therapy - Cognition (FACT-Cog),^21^ especially the Perceived Cognitive Impairment (PCI) subscale that contains 20 items and yields a maximum score of 72. Clinically significant symptoms of cognitive complaints were defined as ratings lower than the 10th percentile of a normative sample.^22^

#### Assessment of lockdown experience

Patients completed a questionnaire about their living conditions and feelings during the first lockdown, including their family environment, education, occupational status, fears about COVID-19 infection, social interactions, feeling of social isolation, and use of psychotropic drugs.

### Statistical analysis

Data were described by using number and proportion for categorical variables, mean and standard deviation for continuous variables. We used a logistic mixed model to describe the evolution of moderate-to-severe PTSD symptoms (IES-R ≥33) over time. Similarly, we performed a linear mixed model to study the IES-R as a continuous measure. Models included an intercept representing baseline symptoms, a discrete time representing change of symptoms over time, and a random intercept for each patient to account for inter-individual variability. To explore associations of factors with PTSD symptoms at baseline and their change over time, factors were entered individually as both simple effect and in interaction with time and were retained in the final multivariable model if p-value<0.10. Models were adjusted for study center and progressive disease at 6 months. In secondary analyses, we used mixed models to describe the evolution of QoL, cognitive complaints and insomnia over time according to PTSD symptoms (IES-R ≥33). In supplementary analyses, we identified distinct trajectories of IES-R using latent class mixed models, which account for heterogeneity in patterns of change. The optimal number of latent classes from 2 to 6 was chosen according to the Bayesian information criterion. The best statistical fit was reached with four trajectory groups, including two small trajectory groups of similar shape which were pooled *a posteriori* (Supplementary Table 1 and Supplementary Figure 1). Multinomial logistic regression was then fitted to identify factors associated with each latent class trajectory.

All statistical analyses were carried out using R statistical software (4.1.0) using package lme4 (version 1.1-27.1), and lcmm (version 1.9.3). A two-side p-value <0.05 was considered statistically significant.

### Ethical Approval

Approval for the study was obtained from the local ethics committee (ref. 220 C07; South Mediterranean II Committee for the Protection of Persons). The study was conducted in compliance with the French research standard (MR-003 “Research in the Field of Health Without Collection of Consent”; compliance commitment to MR-003 for the François Baclesse Center [no. 2146328 v.0, dated from January 26, 2018]). All patients received information, and none expressed any opposition to the use of their data. The trial is registered as ID RCB: 2020-A00879-30, ClinicalTrials NCT04366154.

## RESULTS

### Baseline characteristics

Among the 565 patients who filled in the IES-R questionnaire at baseline, 386 completed at least one follow-up assessment and were included in the present longitudinal analysis (Figure 1). Mean follow-up time was 9.7 months (range 1.8-14.4). Patients were mostly female (76%), had breast cancer (50%), had a good general status (95% of EGOG status 0 or 1), and half of them had metastatic disease (Table 1). The clinical characteristics of patients were similar between the 386 patients retained in the longitudinal analysis and the initial population of 565 patients (Supplementary Table 2). The IES-R score was also comparable between groups at baseline (mean [SD] = 21.5 [15.5] and 20.8 [15.9] respectively). There were 15% of patients living alone, 44% who reported feeling socially isolated and 66% who worried about COVID-19 infection (Table 1).

**Figure 1.**
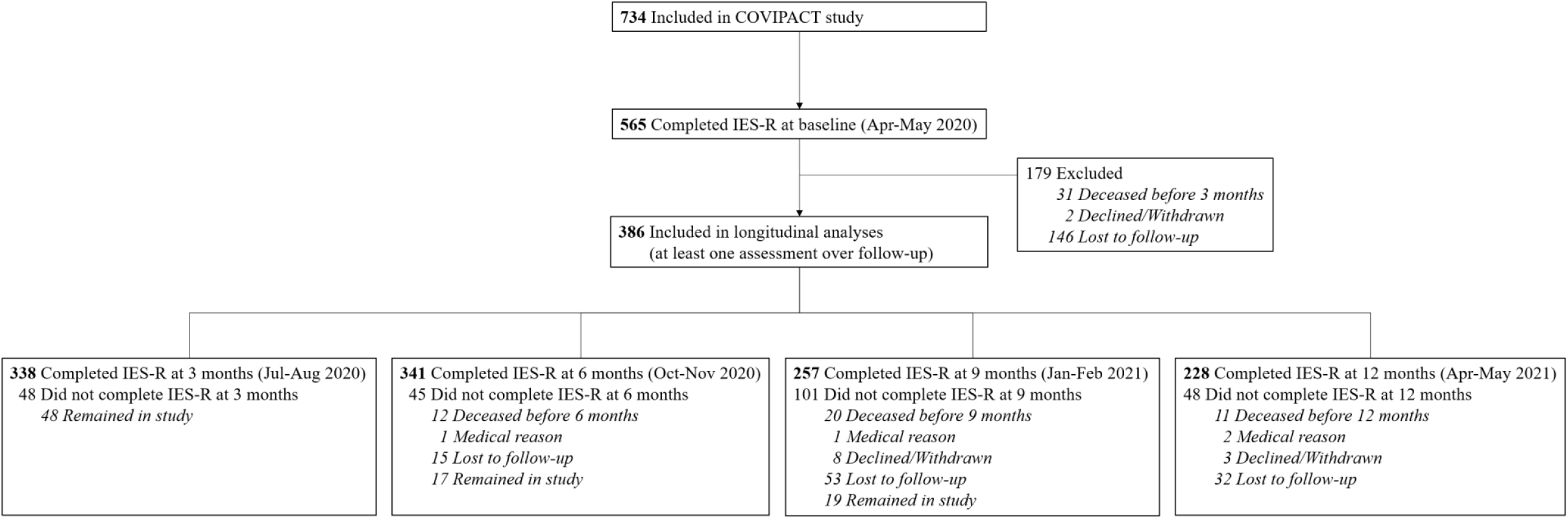
Flow chart of cancer patients in COVIPACT study.

**Table 1.** Baseline characteristics of patients.

| Characteristics | Total<br>(N=386) |
| --- | --- |
| **Clinical characteristics** |  |
| Age, median (min-max) | 63 (28-87) |
| Age, No. (%) |  |
| <70 | 287 (74%) |
| ≥70 | 99 (26%) |
| Sex, No. (%) |  |
| Male | 94 (24%) |
| Female | 292 (76%) |
| Study center, No. (%) |  |
| François Baclesse center | 296 (77%) |
| Henri Becquerel center | 90 (23%) |
| ECOG performance status, No. (%) |  |
| 0 or 1 | 366 (95%) |
| >1 | 20 (5%) |
| Months since diagnosis, median (min-max) | 14 (0.7-410) |
| Type of cancer, No. (%) |  |
| Breast cancer | 193 (50%) |
| Digestive cancer | 44 (11%) |
| Lung, head and neck cancer | 64 (17%) |
| Urologic and gynecologic cancer | 71 (18%) |
| Other solid and hematologic cancer | 14 (4%) |
| Metastatic cancer,<sup>a</sup> No. (%) |  |
| Yes | 204 (54%) |
| No | 173 (46%) |
| History of anxiety-depression, No. (%) |  |
| Yes | 32 (8%) |
| No | 354 (92%) |
| Adapted cancer treatment or care during first lockdown, No. (%) |  |
| Yes | 112 (29%) |
| No | 274 (71%) |
| **Self-reported characteristics** |  |
| Level of education,<sup>b</sup> No. (%) |  |
| Less than high school | 189 (52%) |
| High school diploma | 57 (16%) |
| Post-secondary education | 117 (32%) |
| Current occupation status,<sup>c</sup> No. (%) |  |
| In activity | 95 (26%) |
| Retired | 189 (51%) |
| Not active | 86 (23%) |
| COVID-19-induced financial consequences,<sup>d</sup> No. (%) |  |
| Moderate/Severe | 31 (8%) |
| Little | 43 (12%) |
| None | 291 (80%) |
| Living alone during first lockdown,<sup>e</sup> No. (%) |  |
| Yes | 56 (15%) |
| No | 316 (85%) |
| Feeling of social isolation during first lockdown,<sup>f</sup> No. (%) |  |
| Moderate/Severe | 164 (44%) |
| Not much | 206 (56%) |
| Fear of COVID-19 infection during first lockdown,<sup>g</sup> No. (%) |  |
| Moderate/Severe | 245 (66%) |
| Not much | 126 (34%) |
| Increased use of psychotropic drugs during first lockdown,<sup>h</sup> No. (%) |  |
| Yes | 94 (26%) |
| No | 267 (74%) |

<sup>a</sup> Metastatic cancer not available for 9 hematologic cancer patients. % are of nonmissing data.
<sup>b</sup> Level of education not provided by 23 patients. % are of nonmissing data.
<sup>c</sup> Current occupation status not provided by 16 patients. % are of nonmissing data.
<sup>d</sup> COVID-19-induced financial consequences not provided by 21 patients. % are of nonmissing data.
<sup>e</sup> Living alone during first lockdown not provided by 21 patients. % are of nonmissing data.
<sup>f</sup> Feeling of social isolation during first lockdown not provided by 16 patients. % are of nonmissing data.
<sup>g</sup> Fear of COVID-19 infection during first lockdown not provided by 15 patients. % are of nonmissing data.
<sup>h</sup> Increased use of psychotropic drugs during first lockdown not provided by 25 patients. % are of nonmissing data.

### Evolution of PTSD symptoms over time

At baseline during the first lockdown, 21.5% of patients had moderate-to-severe PTSD symptoms (Figure 2). The rate changed significantly over time (p<0.001) with a marked decrease during the lockdown release (13.6% at M3), followed by a return to the same level as during the first lockdown during the second lockdown (23.2% at M6). The rate of patients with PTSD symptoms remained stable after the second lockdown (22.7% at M9) and slightly decreased at the end of follow-up (17.5% at M12). We observed a similar trend when using the IES-R scale as a continuous measure. All the three PTSD components (avoidance, intrusion, and hyperarousal) followed a similar trajectory over time (data not shown).

**Figure 2.**
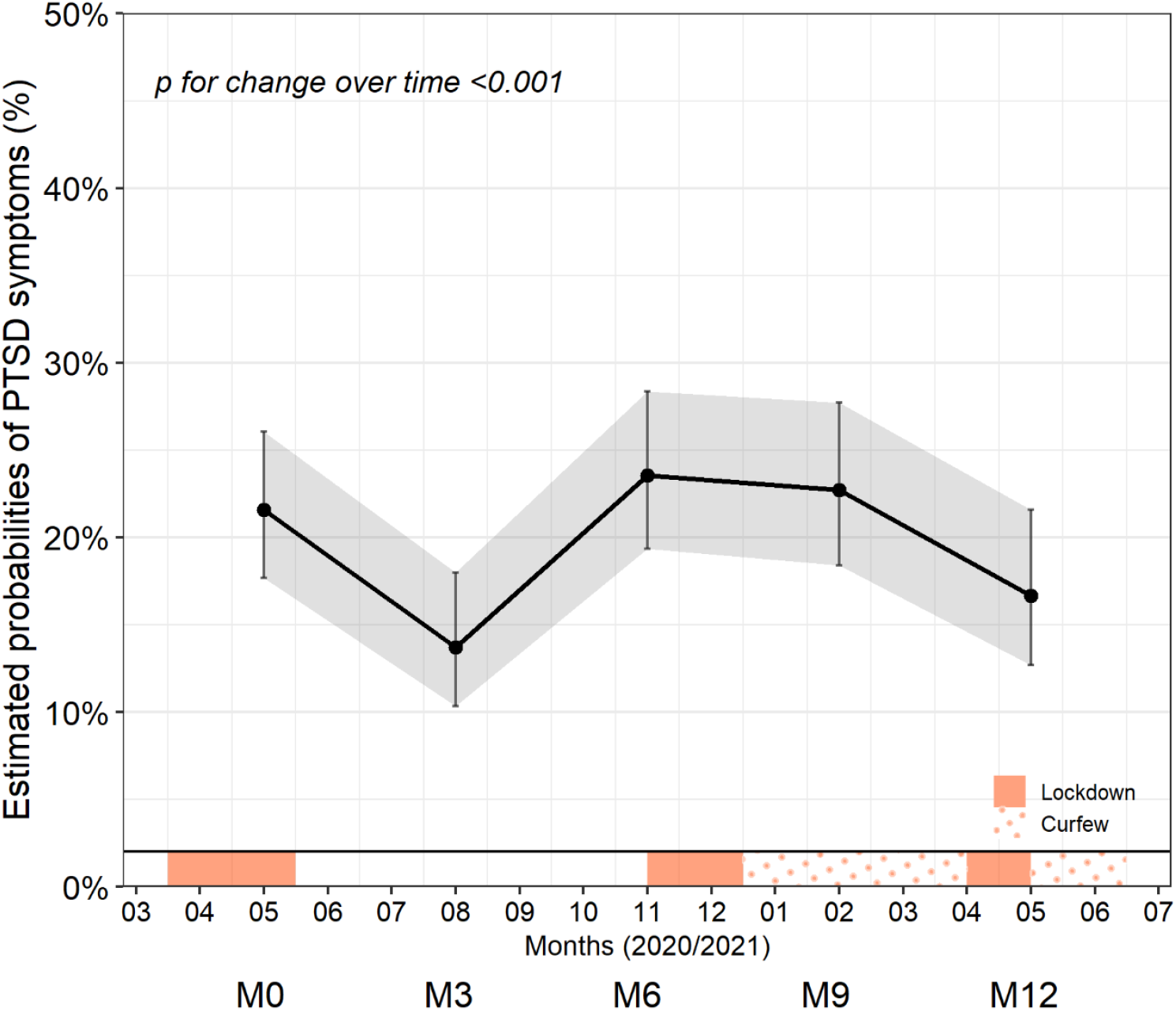
Trajectory of post-traumatic stress disorder (PTSD) symptoms during COVID-19 pandemic in cancer patients in French COVIPACT study (N=386) Estimates are from a logistic mixed model on PTSD symptoms (defined as IES-R score ≥33) with discrete time and a random patient effect.

Female patients, patients who felt socially isolated during the first lockdown, patients who had fears of COVID-19 infection, and those who increased their use of psychotropic drugs during this period had more PTSD symptoms at baseline and these associations remained constant over time (Figure 3). In addition, patients who experienced any adjustment in oncology practices during the first lockdown tended to have more PTSD symptoms at baseline (p=0.053), but the difference decreased and disappeared during follow-up (p for change=0.002). All these associations remained similar when considered simultaneously in a multivariable model (Supplementary Table 3).

**Figure 3.**
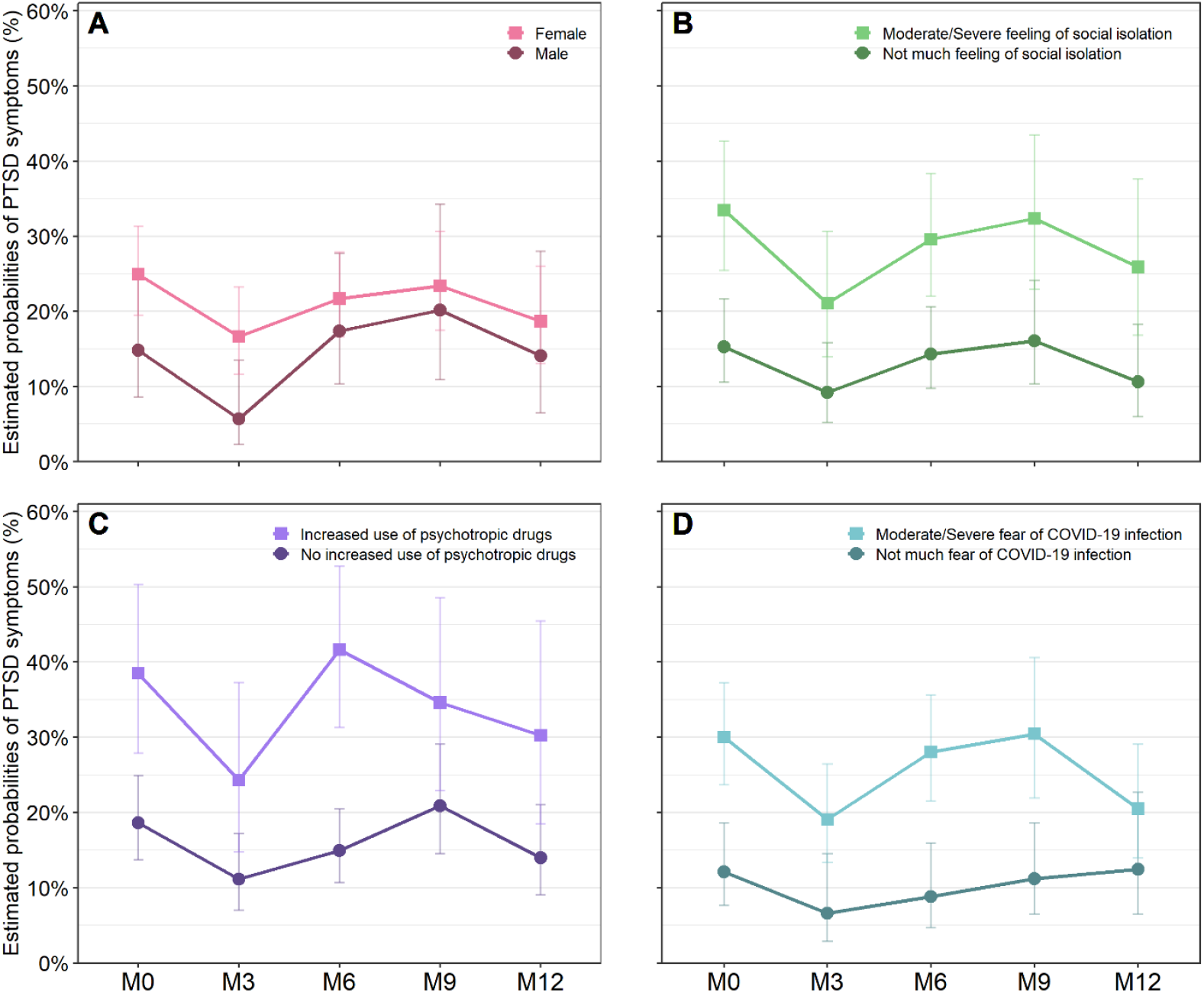
Trajectories of post-traumatic stress disorder (PTSD) symptoms during COVID-19 pandemic by characteristics of cancer patients in COVIPACT study (N=386). Estimates are from separate logistic mixed models on PTSD symptoms (defined as IES-R score ≥33), with discrete time and random patient effect and including as independent fixed factor: patient sex (A), feeling of social isolation (B), increased use of psychotropic drugs (C) and fear of COVID-19 infection during first COVID-19 lockdown in France (D).

We grouped patients into three homogeneous trajectory classes of IES-R change over time (Figure 4). Most patients were in the “Stable low IES-R” class (n=295; 76.4%) characterized by a low IES-R score throughout follow-up. Other patients had a persistently moderate IES-R score over time with an either subtle decrease during follow-up (“Decreased moderate IES-R”, n=23; 6%) or a steep increase at second lockdown (“Increased moderate IES-R”; n=68; 17.6%). As observed in the primary analysis, sex, feeling of social isolation, fear of COVID-19 infection and use of psychotropic drugs were associated with the class trajectories (Supplementary Table 4).

**Figure 4.**
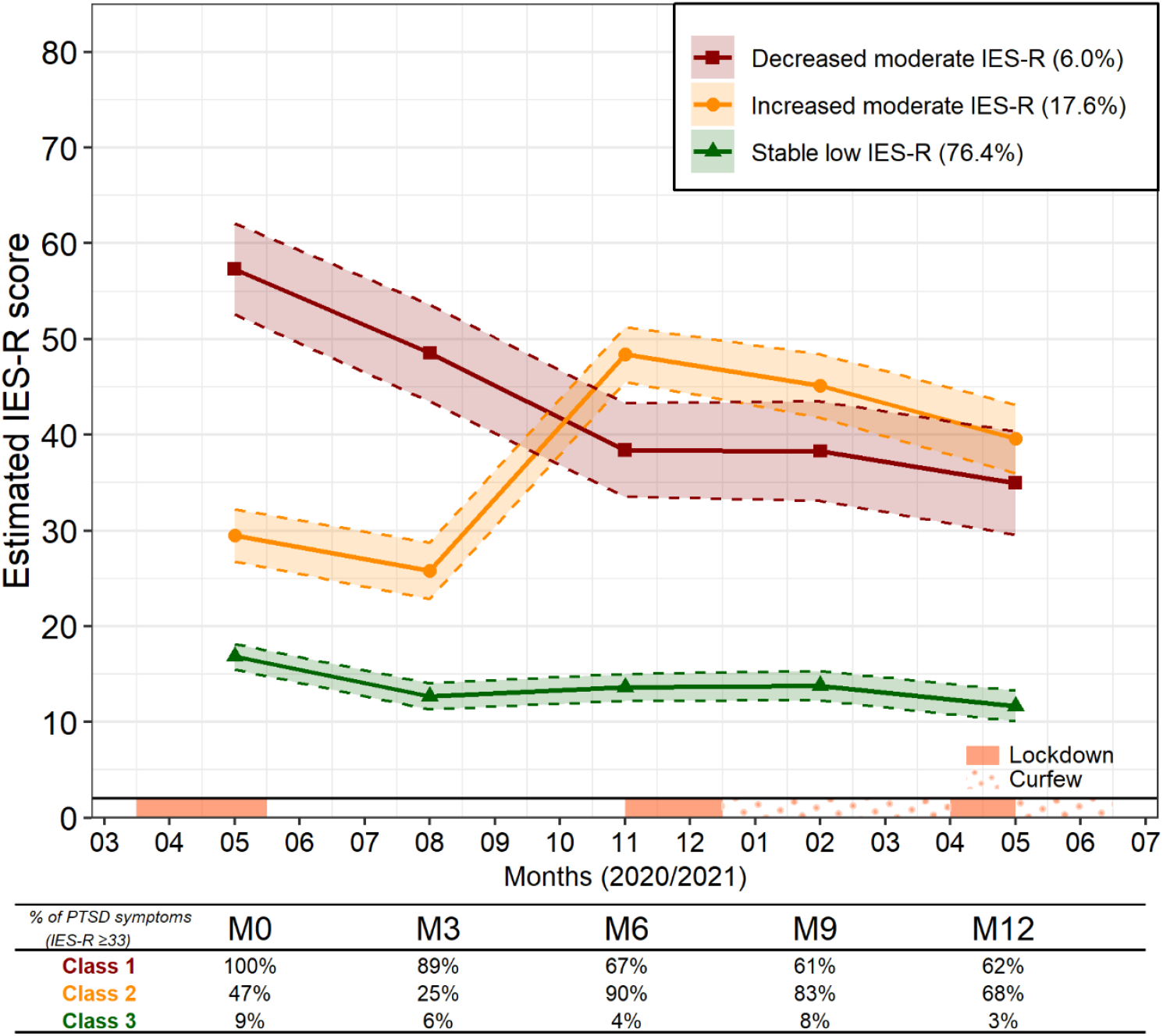
Distinct trajectories of IES-R change over time in cancer patients in French COVIPACT study (N=386). Trajectory classes were identified by latent class linear mixed model on IES-R score. Optimal number of latent classes from 2 to 6 was chosen according to Bayesian information criterion. Four trajectory groups were identified, including one with small sample size (n=5) which was combined a posteriori with another group with a similar trajectory, yielding three trajectory groups with distinct IES-R change over time.

### PTSD-related symptoms

At all times, PTSD symptoms were strongly associated with more insomnia, more cognitive complaints and poor global QoL score (all p<0.001; Figure 5). In particular, 55% of the patients with PTSD symptoms at M6 reported moderate-to-severe insomnia and 40% had cognitive complaints, compared to 25% and 23% of patients without PTSD symptoms, respectively. In addition, there was an average difference of 8 points on the FACT-G global score of QoL throughout follow-up between patients with and without PTSD symptoms.

**Figure 5.**
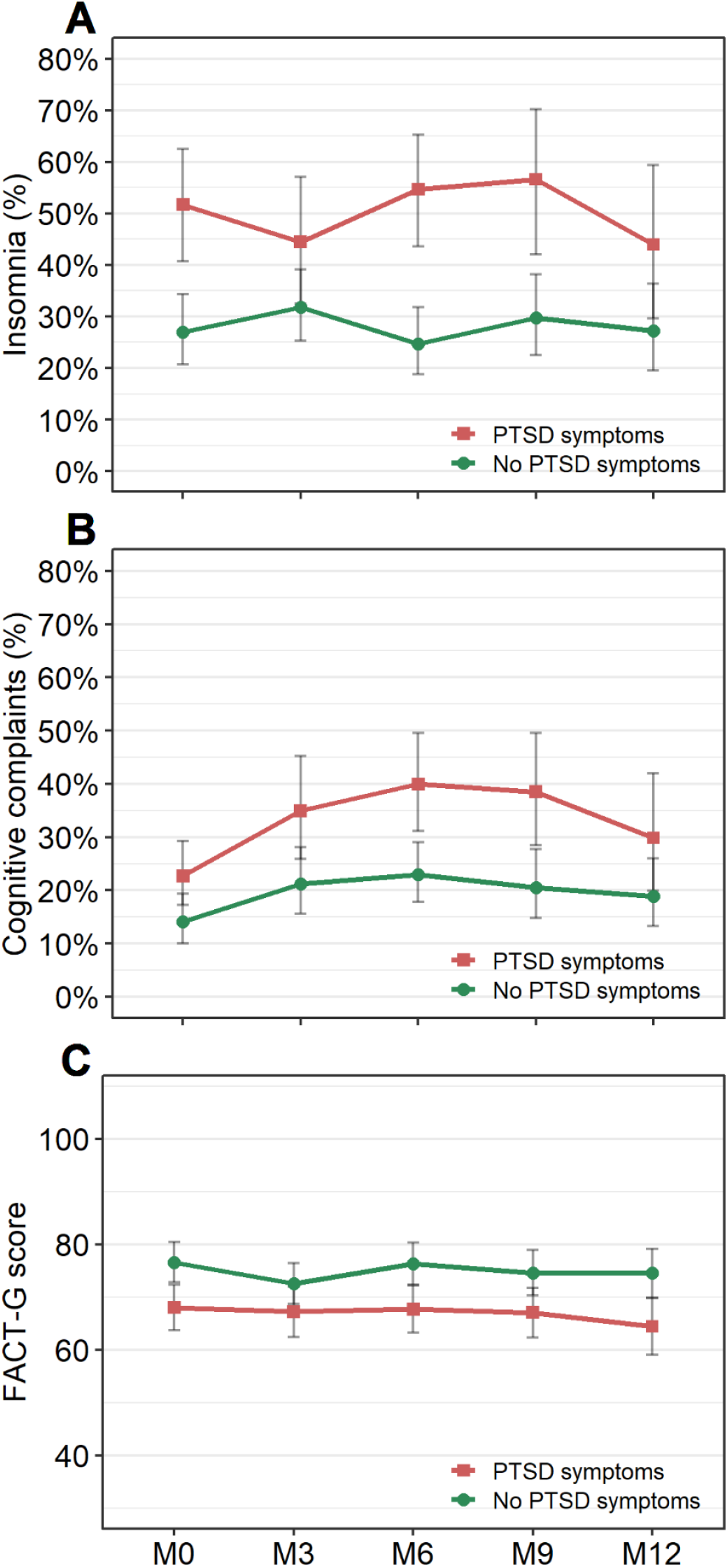
Changes in insomnia, cognitive complaints and quality of life (FACT-G) by post-traumatic stress disorder (PTSD) symptoms during COVID-19 pandemic in cancer patients in French COVIPACT study. Estimates are from logistic and linear mixed models adjusted for baseline age, sex, study center and cancer progression at M6. Curves are plotted for an average study participant profile (a female patient from the François Baclesse center aged <70 years who did not progress at M6). PTSD symptoms are defined as IES-R score ≥33. Insomnia is defined as ISI score ≥15. Cognitive complaints are defined as FACT-Cog PCI score ≥age-dependent cut-off (59 for patients aged under 50y, 47 for patients aged 50-69y, 41 for patients older than 70y). Quality of life is measured using the total score of the FACT-G, with higher scores indicating better quality of life.

## DISCUSSION

This longitudinal prospective study of French cancer patients followed over one year during the COVID-19 pandemic showed high levels of PTSD symptoms over the repeated waves of infections and associated lockdown periods, affecting almost one quarter of patients. Females patients, patients who felt socially isolated during the first lockdown, patients who had fears of COVID-19 infection, and those with increased use of psychotropic drugs during this period presented more PTSD symptoms. PTSD symptoms were associated with more insomnia, more cognitive complaints and worse QoL.

We observed 21.5% of moderate-to-severe PTSD symptoms at baseline at the beginning of the pandemic, which is similar to results of an Italian study of lymphoma patients treated during the same period,^13^ with epidemic conditions equivalent to those in France. A higher rate (35%) was observed in breast cancer patients in Wuhan (China), which may be explained by the exclusively female population and the study location at the COVID-19 epicenter.^14^ Overall, the rate of PTSD symptoms in the cancer population appeared higher during this period compared to what is generally observed in non-pandemic contexts (6 to 9%). ^23,24^

Our results suggest that PTSD symptoms decreased in cancer patients during the first lockdown release (summer 2020), affecting 13.6% of patients. This is in line with another French survey that identified 14.7% of cancer patients with moderate-to-severe PTSD symptoms during the same release period.^25^ Our results are also consistent with data in general population samples showing a decline in peritraumatic distress and anxiety from the early stages of lockdown to the release period.^7,26^ We also observed that the rate of PTSD symptoms increased again at second lockdown, suggesting a strong link between the psychological distress of individuals and the series of restrictions and release periods. While no longitudinal data are available on PTSD changes over time during the COVID-19 pandemic in cancer patients, this further increase in symptoms was recently reported in a UK representative population survey.^27^ However, we did not observe the same trend at the third lockdown, which could be explained by the advent of vaccination and the ability of patients to cope better.

In our study, we found that female sex, feeling socially isolated during the first lockdown, having fears of COVID-19 infection and an increased use of psychotropic drugs were associated with PTSD symptoms. Female sex is a known risk factor for PTSD,^3^ and this susceptibility has also been reported in the context of the COVID-19 pandemic in cancer and general populations.^7,13^ Moreover, loneliness and worry about being infected with COVID-19 have been described as core features of the syndrome triggered by the pandemic. ^28,29^ In addition, patients with PTSD symptoms had poor QoL and more cognitive and insomnia complaints, all of which are known to be associated with PTSD.^3,24^ In the context of the COVID-19 pandemic, sleep disturbances and impaired physical QoL have also been related to higher psychological distress.^30,31^

We identified three distinct trajectories of IES-R change over time. While most patients had a stable low level of PTSD symptoms during the first year of the pandemic, almost a quarter had moderate distress throughout the period, and in particular, 17.6% of patients showed worsened symptoms during the second lockdown. Mental health trajectories in the general population have already been investigated.^26,32^ A 6-week Chinese study conducted at the beginning of the pandemic found that half of the participants were resilient, 22% had delayed PTSD, 19% had recovered PTSD and 15% had chronic PTSD.^32^ A UK longitudinal national survey found that around three quarters of individuals had consistently good mental health during the first six months of the pandemic, whereas the others had consistent or worsened psychological distress at a far higher level than before the pandemic.^26^ Altogether, these results suggest that some psychological symptoms may be delayed in a substantial proportion of individuals, possibly due to the repetition of pandemic waves and associated social restrictions.

The COVIPACT study is the first longitudinal study with a long follow-up of one year assessing PTSD symptoms and QoL in cancer patients during the COVID-19 pandemic. We used validated questionnaires at different periods of lockdowns, including the IES-R which is a widely used screening instrument for PTSD symptoms, even though it does not allow a formal diagnosis to be made. PTSD requires exposure to a specific traumatic event, and the question whether the COVID-19 pandemic can be classified as a “sudden” and “catastrophic” trauma has been debated. For this reason, some authors consider pandemic-induced PTSD symptoms as adjustment disorders.^33^ As we had no control group from the general population, we could not evaluate whether cancer patients presented a particular prevalence or evolution of PTSD symptoms over the COVID-19 pandemic. However, our data are consistent with prevalence estimates from a meta-analysis in the general population,^5^ and longitudinal trends over the repeated pandemic waves.^7,26,27^

Overall, we identified a group of cancer patients with persistent PTSD symptoms over the first year of the COVID-19 pandemic who may need psychological support. Long-term follow-up studies should throw light on the consequences of such psychological distress, especially in the field of oncology where distress is correlated with poor adherence to treatment and survival.^34^

## Data Availability

The data that support the findings of this study are available upon reasonable request to the corresponding author.

## NOTES

### Author Contributions

Conceptualization: BC, FJ; Formal analysis: SLA and JL; Funding acquisition: BC; Investigation: EB, FG, GB, ML, MF, LT, DR, FQ; Methodology: SLA, JL, BC; Project administration: JMG, DR, AL; Resources: OR, AM, FJ, ML, AF; Supervision: BG, LFP, BC, FJ; Writing – original draft: EB, SLA, JL, BC, FJ; Writing – review and editing: All authors.

## Acknowledgements

We acknowledge the Northwest Data Center (CTD-CNO) for managing the data. It is supported by grants from the French National League Against Cancer (LNC) and the French National Cancer Institute (INCa). We thank the Clinical Research Associates Chantal Rieux, Bérénice Legrand, Clotilde Pupin, Delphine Bridelance, Loréna Masseline for data collection as well as Bérengère de Gourmont for support in data entry. We acknowledge Ray Cooke for copyediting the manuscript. We also thank all patients who agreed to participate.

