## Supplementary Material for "Post-traumatic stress symptoms in cancer patients during the COVID-19 pandemic: a one-year longitudinal study"

**Supplementary Tables and Figures**

Etienne Bastien,^1^ Sophie Lefèvre‐Arbogast, PhD,^2,3^ Justine Lequesne, PhD,^3^ corresponding author, François Gernier, MSc,^3^ François Cheriffi, PhD,^1^ Olivier Rigal, MD,^4 , 5^ Lydia Guittet, MD, PhD,^2^ Jean‐Michel Grellard, MSc,^3^ Giulia Binarelli, MSc,^2,3^ Marie Lange, PhD,^2,3^ Marie Fernette,^3^ Laure Tron, PhD,^2^ Adeline Morel, MD,^1^ Doriane Richard, PhD,^5^ Bénédicte Griffon, PhD,^3^ Alexandra Leconte, MSc,^3^ Florian Quilan, MSc,^1^ Louis‐Ferdinand Pépin, MD,^5^ Fabrice Jardin, MD, PhD,^5,6^ Marianne Leheurteur, MD,^4^ Audrey Faveyrial, MD,^1^ Bénédicte Clarisse, PharmD, PhD,^3^ and Florence Joly, MD, PhD.^1, 2, 3^

**Affiliations**

1 Medical Oncology Department, François Baclesse Center, Caen France,

2 Anticipe (Interdisciplinary Research Unit for the Prevention and Treatment of Cancer), INSERM Unit 1086, Caen France,

3 Clinical Research, François Baclesse Center, Caen France,

4 Medical Oncology, Henri Becquerel Center, Rouen France,

5 Clinical Research, Henri Becquerel Center, Rouen France,

6 Hematology, Henri Becquerel Center, Rouen France

**Supplementary Table 1: Fit of latent class models identifying distinct trajectory classes of IES-R evolution**

| Model with n classes | loglik | AIC | BIC | SABIC | entropy | Class 1,  N | Class 2,  N | Class 3,  N | Class 4,  N | Class 5,  N | Class 6,  N |
| --- | --- | --- | --- | --- | --- | --- | --- | --- | --- | --- | --- |
| 1 | -6044.58 | 12103.16 | 12130.85 | 12108.64 | 1 | 386 |  |  |  |  |  |
| 2 | -5972.70 | 11971.39 | 12022.82 | 11981.57 | 0.8959 | 358 | 28 |  |  |  |  |
| 3 | -5945.90 | 11929.80 | 12004.96 | 11944.68 | 0.8672 | 21 | 327 | 38 |  |  |  |
| 4 | -5912.71 | 11875.43 | **11974.32** | 11895.00 | 0.8521 | 23 | 63 | 5 | 295 |  |  |
| 5 | -5900.56 | 11863.11 | 11985.74 | 11887.38 | 0.8648 | 36 | 298 | 34 | 5 | 13 |  |
| 6 | -5893.40 | 11860.80 | 12007.16 | 11889.77 | 0.5964 | 9 | 0 | 15 | 14 | 280 | 68 |

**Supplementary Table 2: Clinical characteristics of patients retained in the longitudinal analysis and the initial population available at baseline**

|  | **Sample retained in longitudinal analysis (n=386)** | **Sample available at baseline (n=565)** |
| --- | --- | --- |
| Age, median (min-max) | 63 (28-87) | 63 (24-87) |
| Age, No. (%) |  |  |
| <70 | 287 (74%) | 419 (74%) |
| ≥70 | 99 (26%) | 146 (26%) |
| Sex, No. (%) |  |  |
| Male | 94 (24%) | 155 (27%) |
| Female | 292 (76%) | 410 (73%) |
| Study center, No. (%) |  |  |
| François Baclesse center | 296 (77%) | 465 (82%) |
| Henri Becquerel center | 90 (23%) | 100 (18%) |
| ECOG performance status, No. (%) |  |  |
| 0 or 1 | 366 (95%) | 531 (94%) |
| >1 | 20 (5%) | 34 (6%) |
| Months since diagnosis, median (min-max) | 14 (0.67-410) | 15 (0.48-410) |
| Type of cancer |  |  |
| Breast cancer | 193 (50%) | 252 (45%) |
| Digestive cancer | 44 (11%) | 94 (17%) |
| Lung, head and neck cancer | 64 (17%) | 109 (19%) |
| Urologic and gynecologic cancer | 71 (18%) | 85 (15%) |
| Other solid and hematologic cancer | 14 (4%) | 25 (4%) |
| Metastatic cancer, No. (%) |  |  |
| Yes | 203 (54%) | 328 (59%) |
| No | 174 (46%) | 227 (41%) |
| *Not applicable / Missing* | 9 | 10 |
| History of anxiety-depression |  |  |
| Yes | 32 (8%) | 46 (8%) |
| No | 354 (92%) | 519 (92%) |
| Adapted cancer treatment or care during first lockdown |  |  |
| Yes | 112 (29%) | 149 (26%) |
| No | 274 (71%) | 416 (74%) |

**Supplementary Table 3: Multivariable analysis of post-traumatic stress disorder (PTSD) symptoms (N=357 in complete case analysis)**

|  | **Association at baseline** | | **Association with change over follow-up** | | | | |
| --- | --- | --- | --- | --- | --- | --- | --- |
|  | **β_M0_ (OR)** | **p** | **β_M3_** | **β_M6_** | **β_M9_** | **β_M12_** | **p** |
| **Sex** |  | 0.005 |  |  |  |  |  |
| Male | ref |  |  |  |  |  |  |
| Female | 0.87 (2.40). |  |  |  |  |  |  |
| **Feeling of social isolation during first lockdown** |  | 0.008 |  |  |  |  |  |
| No/not much | ref |  |  |  |  |  |  |
| Moderate/Severe | 0.62 (1.86) |  |  |  |  |  |  |
| **Fear of COVID-19 infection** |  | <0.001 |  |  |  |  |  |
| No/not much | ref |  |  |  |  |  |  |
| Moderate/Severe | 1.25 (3.48) |  |  |  |  |  |  |
| **Increased use of psychotropic drugs during first lockdown** |  | <0.001 |  |  |  |  |  |
| No | ref |  |  |  |  |  |  |
| Yes | 0.87 (2.40) |  |  |  |  |  |  |
| **Current occupation status** |  | 0.13 |  |  |  |  | 0.080 |
| In activity | ref |  |  |  |  |  |  |
| Retired | 0.65 (1.92) |  | -0.21 | 0.10 | -0.13 | 1.41 |  |
| Not active | 0.10 (1.10) |  | -0.11 | 0.85 | 0.80 | 1.69 |  |
| **Adapted cancer treatment or care during first lockdown** |  | 0.17 |  |  |  |  | 0.006 |
| No | ref |  |  |  |  |  |  |
| Yes | 0.42 (1.52) |  | -0.85 | 0.04 | -1.00 | 0.39 |  |

Multivariable logistic mixed model including factors associated with baseline PTSD symptoms or change with p<0.10 in univariable analysis and adjusted for study center and progressive disease at M6.

- If the factor was associated with PTSD at baseline but not with PTSD change at p<0.10, only the association at baseline was included. In this case, the factor is associated at baseline and consistently over the follow-up, i.e., with same trajectory of change.
- If the factor was associated with PTSD change at p<0.10 but not baseline PTSD symptoms, both the association with change and the association at baseline were included in the model. In this case, there is no association/difference at baseline, but there are different trajectories of change over follow-up according to this factor.
- If the factor was associated with both baseline PTSD symptoms and PTSD change at p<0.10, both the association at baseline and the association with change were included in the model. In this case, there is an association/difference at baseline, and there are different trajectories of change over follow-up, i.e. the baseline association either decrease (and possibly disappear) or increase over follow-up.

**Supplementary Table 4: Multivariable associations between patient’s characteristics and post-traumatic stress symptoms trajectory classes**

|  | **RRR Class 2 vs Class 1**  **(Increased moderate IES-R**  **vs Stable low IES-R)** | **RRR Class 3 vs Class 1**  **(Decreased moderate IES-R**  **vs Stable low IES-R)** | **p** |
| --- | --- | --- | --- |
| **Sex** |  |  | 0.045 |
| Male | ref | ref |  |
| Female | 1.06 | 7.48 |  |
| **Months from diagnostic (for 24-mo increase)** | 1.06 | 1.09 | 0.24 |
| **Current occupation status** |  |  | 0.48 |
| In activity | ref | ref |  |
| Retired | 1.36 | 1.24 |  |
| Not active | 2.17 | 1.48 |  |
| **Feeling of social isolation during first lockdown** |  |  | 0.032 |
| No/not much | ref | ref |  |
| Moderate/Severe | 2.14 | 1.93 |  |
| **Fear of COVID-19 infection** |  |  | <0.001 |
| No/not much | ref | ref |  |
| Moderate/Severe | 2.64 | 12.69 |  |
| **Increased use of psychotropic drugs during first lockdown** |  |  | 0.011 |
| No | ref | ref |  |
| Yes | 1.75 | 3.93 |  |

Relative risk ratio (RRR) are from multivariable multinomial logistic regression adjusted for study center and progressive disease at M6 and including factors with p<0.10 in univariate analysis

**Supplementary Figure 1: Four trajectory classes of IES-R change over time**

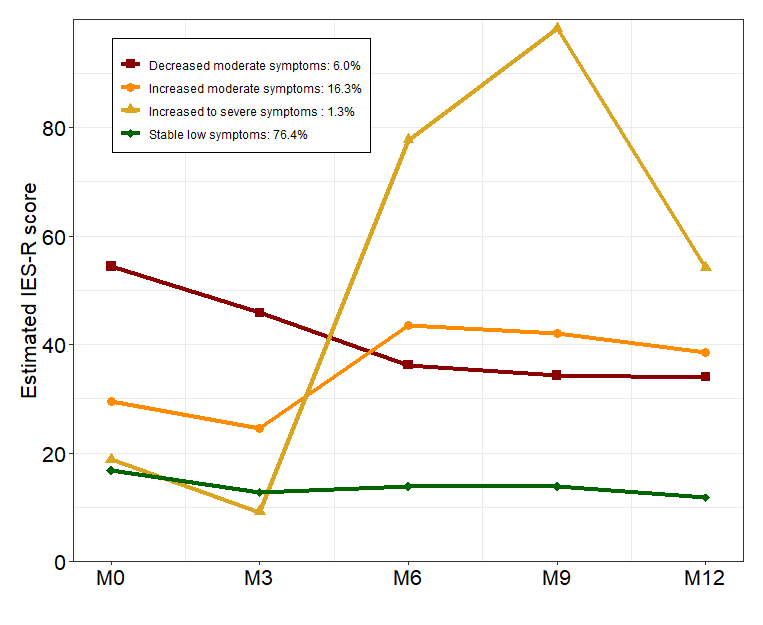

We identified distinct trajectories of IES-R using latent class mixed models, which account for heterogeneity in patterns of change. The optimal number of latent classes from 2 to 6 was chosen according to the Bayesian information criterion. The latent class model that provided best statistical fit identified four trajectory groups. The smallest trajectory class colored in yellow (n=5) was combined a posteriori with the trajectory class colored in orange (n=63) as the “Increased moderate IES-R” group.
